# Factors Associated with Diet Quality in Middle-Aged and Older Adults with HIV: Insights from the PROSPER-HIV study

**DOI:** 10.64898/2026.04.16.26351056

**Authors:** Mari Katundu, Allison R. Webel, Andre P. dos Santos, John D. Cleveland, Dustin M. Long, Vitor H. F. Oliveira, Christine Horvat Davey, Heidi M. Crane, Stephanie A. Ruderman, Thomas W. Buford, Julia Fleming, Kenneth H. Mayer, Greer Burkholder, Barbara Gripshover, Michael S. Saag, Amanda L. Willig

**Affiliations:** University of Alabama at Birmingham, Birmingham, AL; University of Washington, Seattle, WA; Case Western Reserve University, Cleveland, OH; Fenway Health, Boston, MA; Birmingham/Atlanta VA GRECC, Birmingham VA Medical Center, Birmingham, AL; Wake Forest University, Winston-Salem, NC; TW Education LLC, Birmingham, AL

**Keywords:** Acquired Immunodeficiency Syndrome, HIV, Adult, Healthy Diet, Risk Factors, Aging

## Abstract

**Objective:** Improved diet quality is increasingly important for comorbidities management and healthy aging in people with HIV (PWH). Yet, limited data exist on dietary patterns and their correlates in this population. This study aimed to (1) characterize dietary patterns among adult PWH and (2) identify demographic, clinical, and HIV-related factors associated with diet quality.

**Methods:** We conducted a cross-sectional analysis of PWH enrolled in the PROSPER-HIV study across four U.S. academic medical centers. Dietary intake was assessed using three 24-hour dietary recalls and scored using the Healthy Eating Index-2015 (HEI-2015). Participants were categorized into tertiles based on total HEI-2015 scores. Between-group comparisons were performed using Kruskal–Wallis and chi-square tests. Factors independently associated with HEI-2015 scores were identified using multivariable linear regression.

**Results:** A total of 491 PWH were included with a median age of 54 years; 76.4% were male. Overall diet quality was low with inadequate intake of dietary protein, fiber, and micronutrients. When classified by tertiles of HEI-2015 score, higher diet quality was characterized by greater intake of fiber, protein, and key micronutrients. Older age was independently associated with higher HEI-2015 scores (β = 0.119, *p* = 0.018), while Black race was associated with lower scores (β = –3.427, *p* = 0.008). Full-time employment and absence of current pain were marginally associated with better diet quality.

**Conclusions:** Diet quality among PWH varies substantially and is influenced by age, race, and social determinants. Tailored nutritional strategies are needed to support healthy aging and reduce disparities in this population.

## Introduction

Diet quality is a key determinant of chronic disease outcomes, yet it remains understudied in people living and aging with HIV (PWH).^[1, 2]^ Advances in antiretroviral therapy (ART) have transformed HIV into a chronic condition and aging PWH face a heightened burden of comorbidities, including cardiovascular disease, diabetes, and frailty.^[3]^ These conditions are exacerbated by chronic inflammation and immune dysregulation, which persist even with viral suppression.^[4]^ As the demographic profile of PWH shifts towards an older population, optimizing modifiable lifestyle factors, such as diet, is critical to mitigating age-related health disparities in this population.^[5, 6]^

The Healthy Eating Index-2015 (HEI-2015) is a validated tool that assesses adherence to the Dietary Guidelines for Americans, with higher scores linked to reduced morbidity and mortality in the general population.^[7]^ For PWH, poor diet quality is associated with worse metabolic health, immune dysfunction, and accelerated aging.^[8]^ Emerging evidence suggests that nutrient-dense diets may attenuate inflammation and improve ART efficacy, underscoring the need to evaluate dietary patterns in this high-risk group.^[9, 10]^ Diet is influenced by a complex interplay of demographic, clinical, and social factors.^[11, 12]^ Structural inequities, such as food insecurity and limited access to healthy foods, disproportionately affect marginalized groups.^[13–15]^ Employment status and insurance type may further shape dietary choices, while symptoms (e.g., pain, fatigue) may impede meal preparation or adherence to healthy eating.^[16, 17]^ Additionally, older age and female sex have been associated with distinct dietary challenges, such as altered nutrient absorption and caregiving responsibilities.^[18, 19]^ These factors highlight the need for a comprehensive analysis of diet quality correlates, especially in PWH.

While demographic and socioeconomic determinants of diet quality have been well-characterized in general aging populations,^[14, 16]^ their specific associations are less explored among PWH. We thus aimed to: (1) comprehensively characterize dietary patterns in middle-aged and older PWH, and (2) identify both HIV-specific and general population factors associated with diet quality in this vulnerable group. Describing the dietary intake of adults aging with HIV and factors that influence dietary patterns is essential to inform tailored nutrition interventions that promote healthier dietary behaviors in this growing population.

## Methods

### Study Design

We conducted a cross-sectional sub-analysis derived from The Impact of Physical Activity Routines and Dietary Intake on the Longitudinal Symptom Experience of People Living with HIV (PROSPER-HIV) Study, which investigates how physical activity and nutrition affect symptom burden in PWH.^[20]^ PROSPER-HIV participants completed one study visit annually over three years. In the current analysis, we assessed factors associated with diet quality using data from the baseline assessment The study design and reporting followed the Strengthening the Reporting of Observational Studies in Epidemiology (STROBE) guidelines to maintain methodological transparency and quality.^[21]^

### Setting and Participants

Data for this study were sourced from the Center for AIDS Research (CFAR) Network of Integrated Clinical Systems (CNICS), a robust research infrastructure that has compiled comprehensive clinical data on PWH in the United States for over two decades.^[22]^ The PROSPER-HIV study was implemented across four academic medical institutions—Case Western Reserve University (Cleveland, OH), University of Alabama at Birmingham (Birmingham, AL), University of Washington (Seattle, WA), and Fenway Health Institute (Boston, MA) — each serving a representative population of PWH reflective of the broader U.S. context. All study activities related to PROSPER-HIV were carried out within the established CNICS framework. Eligibility criteria required participants to: (1) be active in CNICS (i.e., have provided informed consent and completed the most recent patient-reported outcome measures); (2) be aged 18 years or older; (3) be undergoing ART as part of their CNICS clinical care; and (4) have a recorded HIV viral load below 200 copies/mL at the time of enrollment. Exclusion criteria included: (1) lack of a recent completion of the HIV Symptom Index; (2) current pregnancy, breastfeeding, or plans for pregnancy during the study period; (3) absence of reliable access to a telephone or internet for dietary recall assessments; and (4) intentions to relocate outside the study area within 36 months. Following informed consent, the participants also underwent physical function assessments, answered questionnaires, and completed 24-hour dietary recalls. PROSPER-HIV enrollment (i.e. year 1 visit) happened between January 2019 and August 2022.

All PWH provided informed consent prior to study participation. The PROSPER-HIV study received IRB approval through the University of Washington Institutional Review Board (IRB, #00013048), which served as the primary IRB, with additional reliant approvals granted by the IRBs at the three collaborating sites. The PROSPER-HIV study is registered with ClinicalTrials.gov under identifier NCT03790501.

### Data Collection

#### Dietary Intake

##### Dietary Recalls

Three triple-pass, 24-hour dietary recalls were conducted by trained Registered Dietitians (RDs) at the Dahms Clinical Research Unit (DCRU) at Case Western Reserve University via phone within a 30-day window of the PROSPER-HIV study visit. The recalls included two weekdays and one weekend day and were analyzed using the Nutrition Data System for Research (NDSR) software (University of Minnesota)^[23]^ to determine the average daily intake of macro- and micronutrients, as well as the Healthy Eating Index 2015 (HEI-2015) score. This analysis included individuals who completed at least one dietary recall, with over 80% of participants completing all three recalls. Participants whose recall only reported a total energy intake of less than 800 kcal/day were excluded due to concerns of atypical dietary intake or food intake underreporting.

##### HEI-2015 and Nutrition Consumption

The HEI-2015 is a composite measure of diet quality developed by the United States Department of Agriculture (USDA) to assess how closely dietary intake aligns with the recommendations of the 2015–2020 Dietary Guidelines for Americans (DGA-2015).^[7]^ It is designed to evaluate the balance of food groups in the diet of specific populations using a density-based scoring method (e.g., amount of food per 1,000 kcal). HEI-2015 includes 13 dietary components with a potential score range of either 0-5 or 0-10 that are summed for a total score: Nine “adequacy” components recommended for healthy diets include total vegetables (5), greens and beans (5), total fruits (5), whole fruits (5), whole grains (10), dairy (10), total protein foods (5), seafood and plant proteins (5), and fatty acids ratio (10). Four “moderation” components to consume sparingly include sodium (10), refined grains (10), saturated fats (10), and added sugars (10) (21).

Few studies have used the HEI-2015 to assess individual diets, and therefore, a standardized method for interpreting individual scores has not yet been established. The HEI-2015 is scored on a 100-point scale, with scores >80 indicating good adherence, 50–80 suggesting a need for improvement, and <50 reflecting poor adherence to the guidelines.^[7, 24]^

##### HEI-2015 Tertile Subgroups

Most (98%) participants had an HEI-2015 score <80; therefore participants were categorized into tertiles based on their scores on the HEI-2015 to assess varying levels of diet quality.^[7]^ HEI-2015 scores were ranked in ascending order, and cut-off points were established to ensure a balanced distribution across the three groups. The first tertile included individuals with scores ≤39.5, representing the lowest diet quality. The second tertile comprised scores between 39.6 and 56.6, indicating intermediate diet quality. The third tertile included scores ≥56.7, reflecting higher adherence to dietary guidelines. This classification enabled comparative analyses across distinct levels of dietary quality within the study population.

#### Patient Reported Outcomes

Symptom presence and intensity were assessed through the CNICS clinical assessment of patient-reported outcomes (PROs). As described in the PROSPER-HIV study protocol,^[20]^ these self-administered PRO measures were completed by all participants as part of routine clinical care. The comprehensive assessment battery included measures of HIV-related symptoms (HIV Symptom Index),^[25]^ health-related quality of life (EQ-5D),^[26]^ depression (PHQ-9),^[27]^ anxiety (PHQ-5),^[28]^ alcohol use (AUDIT-C),^[29]^ substance use—including illicit opioids, methamphetamine, cocaine/crack, and marijuana—(ASSIST),^[30]^ and ART regimen. Pain, for example, was evaluated as part of the standard PRO questionnaire using a five-point response scale. For analytical purposes, the response “No, I don’t have pain” was categorized as “no pain,” while the remaining four responses—ranging from “Yes, but it does not bother me” to “Yes, it bothers me a lot”—were grouped as “some pain”.^[31]^

#### Anthropometry

Body Mass Index (BMI) was calculated as weight (kg) divided by height squared (m²), using measurements obtained during the clinic visit that coincided with the PROSPER-HIV study visit. Overweight and obesity were defined as a BMI equal to or greater than 25 kg/m² and 30 kg/m², respectively. Waist and hip circumferences were measured in duplicate by a research assistant using a flexible tape measure and recorded to the nearest 0.1 centimeter (cm). A third measurement was taken if the first two differed by more than 2.54 cm. The average of the two or three measurements was used.

#### Food Security Status

Participants completed a two-item food security questionnaire with the following statements: “The food I/we bought just didn’t last, and I/we didn’t have money to get more,” and “I/we couldn’t afford to eat balanced meals.” Response options were: “Never true”, “Sometimes true”, or “Often true”. The total score ranged from 0 to 2. Food security status was categorized as follows: a score of 0 indicated food security, 1 indicated low food security, and 2 indicated very low food security.^[32]^

#### Covariates

Demographic and anthropometric data—including age, sex at birth, race/ethnicity, height, weight, and BMI—served as potential covariates. Additional information on participants’ demographic characteristics, HIV treatment-related variables, comorbidity diagnoses, and metabolic biomarkers was obtained from the CNICS data repository.^[22]^

#### Statistical Analysis

Participants were categorized into tertiles based on total HEI-2015 scores to reflect low, intermediate, and high diet quality within the sample. Descriptive statistics were used to summarize demographic, clinical, and dietary variables. Continuous variables were reported as medians and interquartile ranges (IQR), and categorical variables as frequencies and percentages. Group comparisons across HEI-2015 tertiles were performed using Kruskal–Wallis tests for continuous variables and chi-square tests for categorical variables. A multivariable linear regression model was used to identify factors independently associated with HEI-2015 total scores. Covariates included age, sex, race, food security, insurance type, employment status, site, HIV transmission risk, and pain status. Results are presented as β estimates with standard error (SE), 95% confidence intervals (CI), t-values, and p-values. All data were analyzed with SAS version 9.4 (SAS Institute Inc., Cary, NC, USA) with a significance level of *p*<0.05.

## Results

**Table 1** summarizes the demographic, clinical, and behavioral characteristics of 491 middle-aged and older adults with HIV, stratified by HEI-2015 total score tertiles. Participants in the highest HEI tertile were slightly older (median = 55 years; p = 0.046) and had a significantly lower proportion of females (14.1%) compared to the lowest tertile (28.2%; p = 0.002). Racial distribution was strongly associated with diet quality (*p* < 0.001), with White individuals more prevalent in the highest tertile (54.0%) and Black individuals in the lowest (66.3%). Insurance status also varied significantly (*p* < 0.001), with a higher proportion of participants in the highest tertile reporting unknown insurance (31.9%) and fewer with public insurance (28.2%) compared to the lowest tertile (48.5%). HIV transmission risk differed across tertiles (*p* = 0.050), with heterosexual contact more common in the lowest tertile (32.5%) and men who have sex with men (MSM) predominating in the highest (71.2%). Food security increased with diet quality (*p* = 0.020), from 61.3% in the lowest to 76.7% in the highest tertile. Employment status was positively associated with HEI (p = 0.014), with full-time employment more frequent in the highest tertile (46.0%) than in the lowest (30.1%). No significant differences were observed in at-risk alcohol use (*p* = 0.994) or recent drug use (*p* = 0.175), although both were numerically lower in the highest tertile. Reports of current pain were significantly less frequent in the highest tertile (28.2%) compared to the lower ones (42.3% and 44.8%; p = 0.023), while symptoms of fatigue, anxiety, and depression did not differ. HIV-related clinical markers, including CD4 count and time in care, were similar across groups, though a small difference in viral load was noted (*p* = 0.046). Among clinical biomarkers, fasting glucose (*p* = 0.002) and hemoglobin A1c (*p* = 0.020) were significantly lower in the highest tertile.

**Table 1:** Demographic, Clinical, and Behavioral Characteristics of Middle-Aged and Older Adults with HIV, Stratified by Tertiles of the Healthy Eating Index-2015 (HEI-2015).

| n (%) or median (IQR) | Overall<br>n = 491 | 1<sup>st</sup> HEI tertile<br>n =163 (33.2%) | 2<sup>nd</sup> HEI tertile<br>n = 165 (33.6%) | 3<sup>rd</sup> HEI tertile<br>n = 163 (33.2%) | P |
| --- | --- | --- | --- | --- | --- |
| **Age, years old** | 54 (45; 61) | 53 (43; 59) | 55 (48; 61) | 55 (44; 64) | **0.046** |
| **Birth Sex** |  |  |  |  |  |
| Female | 116 (23.6) | 46 (28.2) | 47 (28.5) | 23 (14.1) | **0.002** |
| Male | 375 (76.4) | 117 (71.8) | 118 (71.5) | 140 (85.9) |  |
| **Race** |  |  |  |  |  |
| Black | 270 (55.0) | 108 (66.3) | 98 (59.4) | 64 (39.3) | **<.001** |
| White | 193 (39.3) | 47 (28.8) | 58 (35.2) | 88 (54.0) |  |
| Other | 28 (5.7) | 8 (4.9) | 9 (5.5) | 11 (6.7) |  |
| **Hispanic, yes** | 29 (5.9) | 6 (3.7) | 9 (5.5) | 14 (8.6) | 0.221 |
| **Anthropometry** |  |  |  |  |  |
| BMI, kg/m<sup>2</sup> | 28.5 (24.9; 33.0) | 28.3 (24.0; 33.3) | 29.0 (25.2; 33.3) | 28.4 (25.5; 32.0) | 0.415 |
| BMI category |  |  |  |  | 0.213 |
| Underweight | 11 (2.2) | 7 (4.3) | 1 (0.6) | 3 (1.8) |  |
| Normal weight | 114 (23.2) | 42 (25.8) | 36 (21.8) | 36 (22.1) |  |
| Overweight | 177 (36.0) | 52 (31.9) | 59 (35.8) | 66 (40.5) |  |
| Obese | 189 (38.5) | 62 (38.0) | 69 (41.8) | 58 (35.6) |  |
| Height, cm | 175.3<br>(167.6; 180.3) | 175.3<br>(167.6; 180.3) | 174.0<br>(167.6; 180.3) | 177.8<br>(170.2; 182.9) | 0.026 |
| Weight, kg | 87.1 (75.4; 99.4) | 85.9 (73.5; 98.3) | 87.1 (75.5; 99.4) | 87.7 (76.2; 100) | 0.481 |
| Waist, cm | 102 (91; 112) | 102 (92; 111) | 101 (92; 113) | 101 (91; 111) | 0.873 |
| Hip, cm | 105 (98; 115) | 106 (99; 117) | 105 (98; 115) | 105 (97; 113) | 0.503 |
| **Food security status** |  |  |  |  |  |
| Food secure | 328 (66.8) | 100 (61.3) | 103 (62.4) | 125 (76.7) | **0.020** |
| Low food secure | 71 (14.5) | 25 (15.3) | 27 (16.4) | 19 (11.7) |  |
| Very low food secure | 92 (18.7) | 38 (23.3) | 35 (21.2) | 19 (11.7) |  |
| **Employment status** |  |  |  |  |  |
| Full-time | 187 (38.1) | 49 (30.1) | 63 (38.2) | 75 (46.0) | **0.014** |
| Part-time | 53 (10.8) | 24 (14.7) | 12 (7.3) | 17 (10.4) |  |
| Unemployed | 251 (51.1) | 90 (55.2) | 90 (54.5) | 71 (43.6) |  |
| **Insurance** |  |  |  |  |  |
| Private | 164 (33.4) | 59 (36.2) | 51 (30.9) | 54 (33.1) | **0.001** |
| Public | 198 (40.3) | 79 (48.5) | 73 (44.2) | 46 (28.2) |  |
| Uninsured | 26 (5.3) | 6 (3.7) | 9 (5.5) | 11 (6.7) |  |
| Unknown | 103 (21.0) | 19 (11.7) | 32 (19.4) | 52 (31.9) |  |
| **HIV transmission risk factor** |  |  |  |  |  |
| Heterosexual contact | 142 (28.9) | 53 (32.5) | 56 (33.9) | 33 (20.2) | 0.050 |
| MSM | 306 (62.3) | 95 (58.3) | 95 (57.6) | 116 (71.2) |  |
| MSM-IDU | 13 (2.6) | 3 (1.8) | 3 (1.8) | 7 (4.3) |  |
| Injection drug use | 14 (2.9) | 5 (3.1) | 7 (4.2) | 2 (1.2) |  |
| Unknown | 16 (3.3) | 7 (4.3) | 4 (2.4) | 5 (3.1) |  |
| **Alcohol use** |  |  |  |  |  |
| At risk drinking | 74 (15.6) | 24 (15.4) | 25 (15.7) | 25 (15.8) | 0.994 |
| Not at risk | 399 (84.4) | 132 (84.6) | 134 (84.3) | 133 (84.2) |  |
| **Drug in last 3 months, yes** |  |  |  |  |  |
| Current use | 36 (7.6) | 14 (8.9) | 15 (9.5) | 7 (4.4) | 0.175 |
| **Symptoms** |  |  |  |  |  |
| Pain, having today | 189 (38.5) | 69 (42.3) | 74 (44.8) | 46 (28.2) | **0.023** |
| Fatigue, having today | 263 (53.6) | 94 (57.7) | 85 (51.5) | 84 (51.5) | 0.438 |
| Anxiety |  |  |  |  | 0.237 |
| Extremely | 13 (2.8) | 8 (5.1) | 3 (1.9) | 2 (1.3) |  |
| Some | 118 (25.2) | 38 (24.2) | 43 (27.6) | 37 (23.7) |  |
| None | 338 (72.1) | 111 (70.7) | 110 (70.5) | 117 (75.0) |  |
| Depression |  |  |  |  | 0.533 |
| Severe | 25 (5.2) | 10 (6.2) | 9 (5.5) | 6 (3.8) |  |
| Mild | 139 (28.8) | 52 (32.3) | 46 (28.2) | 41 (25.8) |  |
| None | 319 (66.0) | 99 (61.5) | 108 (66.3) | 112 (70.4) |  |
| **HIV status** |  |  |  |  |  |
| CD4 count, cells/μl | 695 (480; 933) | 707 (481; 1,000) | 716 (480; 933) | 652 (473; 868) | 0.277 |
| HIV viral load, copies/mL | 20.0 (20.0; 49.0) | 20.0 (20.0; 49.0) | 20.0 (20.0; 49.0) | 20.0 (19.0; 49.0) | **0.046** |
| Time in care, years | 10.0 (6.0; 17.0) | 10.0 (7.0; 16.0) | 10.0 (6.0; 17.0) | 11.0 (6.0; 17.0) | 1.002 |
| **Comorbidities** |  |  |  |  |  |
| Diabetes Mellitus | 102 (20.8) | 33 (20.2) | 36 (21.8) | 33 (20.2) | 0.921 |
| Hypertension | 235 (47.9) | 85 (52.1) | 84 (50.9) | 66 (40.5) | 0.069 |
| **Biomarkers** |  |  |  |  |  |
| Total Cholesterol | 177 (150; 204) | 176 (152; 208) | 177 (152; 202) | 179 (147; 200) | 0.950 |
| LDL | 97 (79; 120) | 99 (80; 121) | 94 (77; 117) | 97 (80; 121) | 0.272 |
| HDL | 48 (40; 58) | 48 (40; 60) | 47 (39; 57) | 48 (40; 58) | 0.836 |
| Triglycerides | 121 (86; 182) | 122 (82; 174) | 131 (90; 205) | 112 (87; 169) | 0.205 |
| Glucose | 88 (77; 99) | 85 (75; 96) | 91 (80; 104) | 89 (79; 98) | **0.002** |
| Hgb A1c | 5.5 (5.2; 5.9) | 5.6 (5.2; 5.9) | 5.7 (5.3; 6.1) | 5.4 (5.2; 5.8) | **0.002** |

Note: BMI = Body Mass Index; MSM = Men Who Have Sex With Men; IDU = Injection Drug Use; ART = Antiretroviral Therapy; NRTIs = Nucleoside Reverse Transcriptase Inhibitors; NNRTIs = Non-Nucleoside Reverse Transcriptase Inhibitors; HDL = High-Density Lipoprotein; LDL = Low-Density Lipoprotein; Hgb A1c = Hemoglobin A1c.

Percentile scores for HEI-2015 components across tertile groups are depicted in **Figure 1**. Participants in the highest HEI tertile exhibited consistently higher scores across all components, with notable differences in total fruit, whole grains, greens and beans, and seafood and plant protein intake, as well as in the fatty acid ratio.

**Figure 1:**
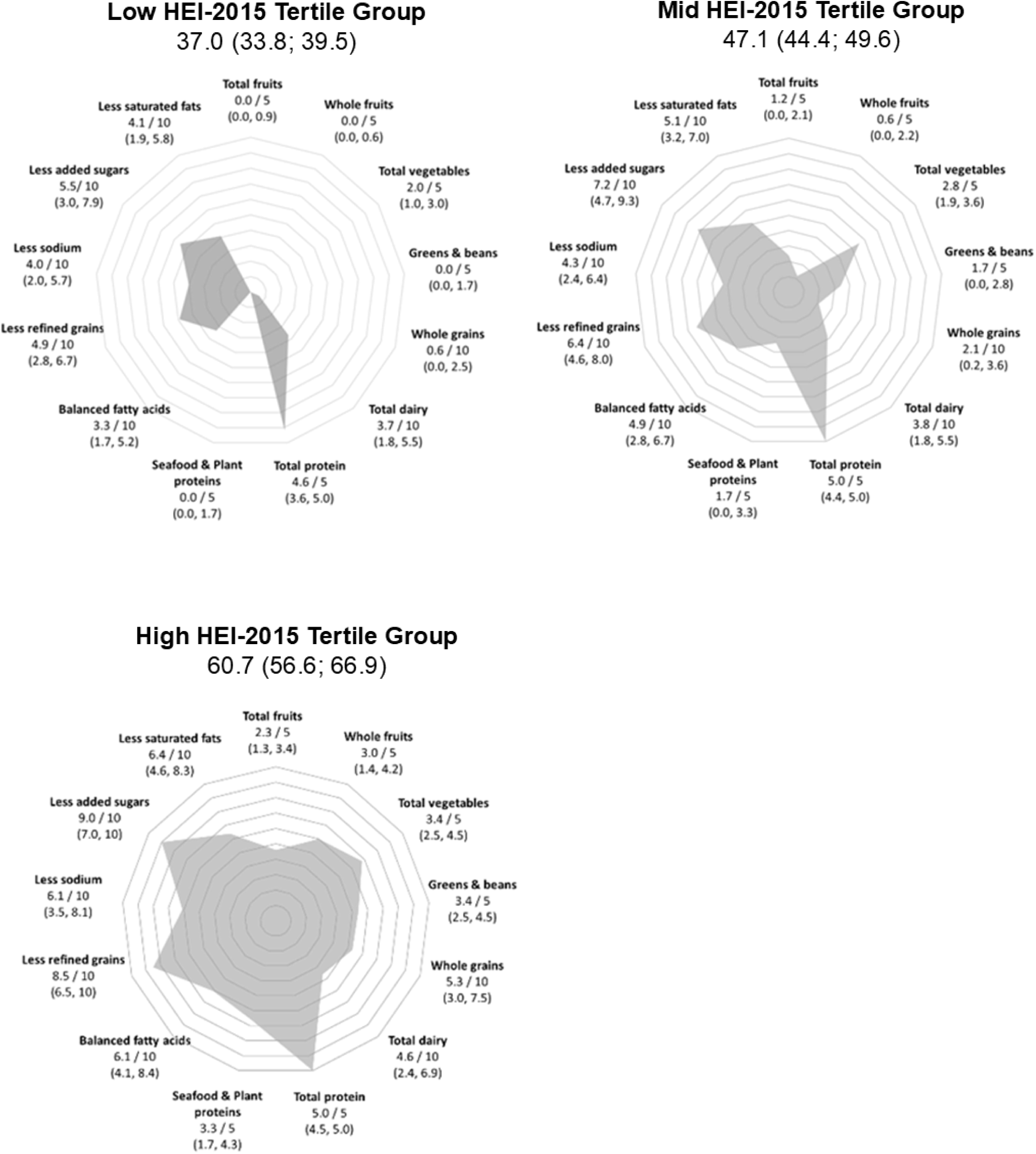
Radar Plots of Healthy Eating Index-2015 (HEI-2015) Component Percentile Scores Across Tertiles of Total HEI-2015 Score. The median score and interquartile range are shown for each of the thirteen dietary components used to compute the total HEI-2015 score.

Self-reported dietary intake across HEI-2015 tertiles is summarized in **Table 2**. Individuals in the highest tertile reported significantly higher protein intake, both in grams per kilogram of body weight *(p* = 0.014) and in total daily grams (*p* = 0.001), as well as a greater proportion of energy from protein (*p* < 0.001). The percentage of energy from saturated fat decreased significantly across tertiles (*p* < 0.001), alongside a reduction in total fat intake (*p* = 0.021), while polyunsaturated fat intake increased (*p* = 0.033). Dietary fiber intake nearly doubled from the lowest to highest tertile (median = 11.000 g vs. 20.200 g; *p* < 0.001), reflecting a key marker of dietary quality and health. Several micronutrients were significantly higher in the highest tertile, including magnesium, potassium (*p* < 0.001 for both), calcium (*p* = 0.032), iron (*p* = 0.007), zinc (*p* = 0.016), vitamin D (*p* = 0.004), vitamin E, vitamin K, vitamin C, and folate (*p* < 0.001 for all). Sodium intake showed a significant inverse association with HEI score (*p* < 0.001), and omega-3 intake was modestly but significantly higher in the top tertile (*p* = 0.009).

**Table 2:** Self-Reported Dietary Intake Among Adults with HIV, Stratified by Tertiles of the Healthy Eating Index-2015 (HEI-2015).

| Variables | Dietary Guideline* | Overall<br>n = 491 | 1<sup>st</sup> HEI tertile<br>n =163 (33.2%) | 2nd HEI tertile<br>n = 165 (33.6%) | 3rd HEI tertile<br>n = 163 (33.2%) | p |
| --- | --- | --- | --- | --- | --- | --- |
| Total HEI-2015 score |  | 47.1 (39.5; 56.6) | 37.0 (33.8; 39.5) | 47.1 (44.4; 49.6) | 60.7 (56.6; 66.9) |  |
| Energy, kcal | - | 1,820.2<br>(1,373.7; 2,307.4) | 1,910.9<br>(1,383.1; 2,458.4) | 1,721.7<br>(1,351.4; 2,188.7) | 1,879.8<br>(1,418.9; 2,250.4) | 0.075 |
| Protein, g/kg* | 1.2 | 0.9 (0.6; 1.2) | 0.8 (0.6; 1.2) | 0.8 (0.6; 1.1) | 0.9 (0.7; 1.2) | 0.014 |
| Total protein, g | - | 75.8 (58.2; 97.0) | 72.7 (55.9; 93.4) | 70.4 (55.5; 91.7) | 86.7 (64.1; 102.6) | 0.001 |
| Protein, energy % | 15-20% | 17.4 (14.3; 20.2) | 16.2 (12.7; 18.6) | 17.6 (14.0; 20.2) | 18.8 (15.9; 21.1) | <.001 |
| Carbo, energy % | 50% | 44.4 (38.6; 50.1) | 45.7 (39.9; 51.2) | 43.4 (37.2; 50.4) | 44.1 (37.5; 48.4) | 0.057 |
| Fat, energy % | 30-35% | 36.7 (31.9; 41.2) | 37.7 (32.9; 42.1) | 36.9 (32.0; 41.9) | 35.6 (30.1; 39.7) | 0.021 |
| SFA, energy % | 10% | 11.9 (9.9; 14.1) | 13.1 (11.3; 15.1) | 12.0 (10.1; 14.0) | 10.6 (8.6; 12.5) | <.001 |
| MUFA, energy % |  | 12.9 (10.8; 15.0) | 13.0 (11.2; 14.7) | 12.8 (10.8; 15.4) | 12.7 (10.4; 15.0) | 0.594 |
| PUFA, energy % |  | 8.1 (6.3; 9.8) | 7.5 (5.8; 9.4) | 8.1 (6.3; 10.0) | 8.4 (6.6; 10.0) | 0.033 |
| Total dietary fiber, g | 28g | 14.2 (9.8; 20.7) | 11.0 (7.6; 15.1) | 13.1 (9.8; 17.9) | 20.2 (14.4; 26.7) | <.001 |
| Calcium, mg | 1300 mg | 705.6<br>(474.7; 953.6) | 706.4<br>(445.9; 910.5) | 662.8<br>(451.2; 916.6) | 719.5<br>(556.7; 1,040.2) | 0.032 |
| Sodium, mg | 2300 mg | 2,862.0<br>(2,146.5; 3,788.3) | 3,304.3<br>(2,298.2; 4,190.3) | 2,727.2<br>(2,154.9; 3,564.5) | 2,621.1<br>(2,003.3; 3,429.4) | <.001 |
| Potassium, mg | 4700 mg | 2,076.0<br>(1,560.5; 2,795.5) | 1,799.6<br>(1,336.2; 2,404.6) | 1,956.7<br>(1,516.1; 2,529.3) | 2,724.6<br>(1,909.7; 3,188.0) | <.001 |
| Magnesium, mg | 420 mg | 233.9<br>(173.0; 311.2) | 193.9<br>(141.0; 263.2) | 213.2<br>(171.4; 284.3) | 295.8<br>(233.7; 388.5) | <.001 |
| Iron, mg | 18 mg | 11.7 (8.4; 15.4) | 11.3 (7.8; 14.8) | 10.9 (7.8; 14.1) | 12.7 (9.2; 17.6) | 0.007 |
| Zinc, mg | 11 mg | 9.5 (7.0; 12.6) | 9.4 (6.5; 12.5) | 8.7 (7.0; 11.8) | 10.2 (7.4; 13.5) | 0.016 |
| Vitamin D, IU | 800 IU | 144 (76; 220) | 132 (64; 204) | 140 (84; 200) | 164 (92; 292) | 0.004 |
| Vitamin E, mcg | 15 mg | 7.1 (4.9; 10.0) | 6.1 (4.1; 9.0) | 6.7 (4.8; 9.4) | 8.6 (6.2; 12.9) | <.001 |
| Vitamin K, mcg | 120 mcg | 82.7 (47.0; 156.1) | 57.9 (37.7; 103.0) | 80.0 (44.4; 127.6) | 119.4 (75.8; 212.2) | <.001 |
| Total folate, mcg | 400 DFE<br>mcg | 303.5<br>(231.2; 426.5) | 281.0<br>(204.0; 407.2) | 282.0<br>(210.6; 413.3) | 359.9<br>(268.5; 467.8) | <.001 |
| Vitamin B12, mcg | 2.4 mcg | 3.6 (2.3; 5.3) | 3.5 (2.0; 5.3) | 3.6 (2.4; 5.0) | 3.8 (2.6; 5.6) | 0.241 |
| Vitamin C, mg | 90 mg | 52.5 (26.0; 89.4) | 33.5 (18.1; 66.5) | 50.0 (25.8; 82.5) | 79.7 (45.0; 130.7) | <.001 |
| Omega 3, g | M1.6g /<br>F1.1g | 1.7 (1.1; 2.4) | 1.6 (1.0; 2.2) | 1.6 (1.1; 2.3) | 1.8 (1.3; 2.8) | 0.009 |
| Alcohol, g | - | 0.0 (0.0; 0.4) | 0.0 (0.0; 0.1) | 0.0 (0.0; 0.2) | 0.0 (0.0; 7.0) | 0.058 |

Note: SFA = Saturated Fatty Acids; MUFA = Monounsaturated Fatty Acids; PUFA = Polyunsaturated Fatty Acids; DFE = Dietary Folate Equivalents; IQR = Interquartile Range. \*Recommended intakes of nutrients vary by age and sex and are known as Recommended Dietary Allowances and Adequate Intakes. However, one value for each nutrient, known as the Daily Value, is used only for reference purposes in this table.

**Table 3** presents the multivariable linear regression results assessing characteristics independently associated with HEI-2015 score. Older age was positively associated with better diet quality (β = 0.119, *p* = 0.018), indicating a trend toward improved dietary adherence in older adults. Race was a significant factor, with Black participants having lower HEI scores compared to White participants (β = −3.427, *p* = 0.008), independent of age, sex, race, food security, insurance type, employment status, site, HIV transmission risk, and pain status. There was a marginal association suggesting higher HEI scores among individuals employed full-time (β = 2.525, *p* = 0.067) and those reporting no current pain (β = 2.117, *p* = 0.066), although these did not reach statistical significance. Other variables, including sex, food security, insurance type, site, HIV transmission risk, and ART regimen, were not significantly associated with diet quality in the adjusted model.

**Table 3:** Multivariable Linear Regression of Factors Associated with Healthy Eating Index-2015 (HEI-2015) Scores.

| Parameter | Estimate | LCI | UCI | SE | t Value | Pr > \|t\| |
| --- | --- | --- | --- | --- | --- | --- |
| Age | 0.119 | 0.02 | 0.22 | 0.05 | 2.38 | **0.018** |
| Birth sex (referent group: Male) |  |  |  |  |  |  |
| Female | -1.899 | -5.64 | 1.85 | 1.91 | -1.00 | 0.319 |
| Race (ref: White) |  |  |  |  |  |  |
| Black | -3.427 | -5.97 | -0.89 | 1.29 | -2.65 | **0.008** |
| Other | -3.078 | -7.78 | 1.63 | 2.39 | -1.29 | 0.199 |
| Food security (ref: Low food security) |  |  |  |  |  |  |
| Food Secure | 0.691 | -2.34 | 3.72 | 1.54 | 0.45 | 0.655 |
| Very low food security | -1.055 | -4.69 | 2.58 | 1.85 | -0.57 | 0.569 |
| Insurance (ref: Private) |  |  |  |  |  |  |
| Public | 0.497 | -2.31 | 3.31 | 1.43 | 0.35 | 0.728 |
| Uninsured | 3.229 | -2.01 | 8.47 | 2.67 | 1.21 | 0.227 |
| Unknown | 3.881 | -0.63 | 8.39 | 2.29 | 1.69 | 0.092 |
| Employment (ref: Unemployed) |  |  |  |  |  |  |
| Full time | 2.525 | -0.17 | 5.22 | 1.37 | 1.84 | **0.067** |
| Part time | 0.516 | -2.96 | 3.99 | 1.77 | 0.29 | 0.771 |
| Site (ref: University of Washington) |  |  |  |  |  |  |
| CWRU | -1.959 | -5.91 | 1.99 | 2.01 | -0.97 | 0.331 |
| FENWAY | 2.817 | -3.54 | 9.17 | 3.23 | 0.87 | 0.384 |
| UAB | -1.990 | -6.47 | 2.49 | 2.28 | -0.87 | 0.384 |
| HIV transmission risk factor (ref: MSM) |  |  |  |  |  |  |
| Heterosexual contact | 1.754 | -1.84 | 5.35 | 1.83 | 0.96 | 0.338 |
| MSM-IDU/IDU/Unknown | 0.298 | -3.74 | 4.33 | 2.05 | 0.15 | 0.885 |
| Pain (ref: Some) |  |  |  |  |  |  |
| Missing data (n=18) | 1.182 | -6.24 | 8.60 | 3.78 | 0.31 | 0.754 |
| None | 2.117 | -0.14 | 4.37 | 1.15 | 1.85 | **0.066** |

Note: CI = Confidence Interval; SE = Standard Error; LCI = Lower Confidence Interval; UCI = Upper Confidence Interval; t Value = t-statistic for the regression coefficient; Pr > |t| = p-value for testing whether the coefficient differs significantly from zero; Ref = Reference category. UW = University of Washington; CWRU = Case Western Reserve University; UAB = University of Alabama at Birmingham.

## Discussion

In this study, we comprehensively described dietary patterns and examined both HIV-specific and general population factors associated with diet quality among middle-aged and older PWH. Adults with HIV reported lower HEI-2015 total scores (median of 47.1) compared to the average population score range of 52-58.^[33]^ Despite this, PWH with higher HEI scores still consumed diets that were more nutrient-dense, with improved macronutrient balance, greater fiber and micronutrient adequacy, and reduced intake of nutrients linked to chronic disease risk. By utilizing validated 24-hour dietary recall methods and stratifying by HEI-2015 tertiles, we captured a nuanced profile of nutrient intake, macronutrient distribution, and food group patterns. These results can inform patient-centered nutritional interventions and policies that aim to reduce health disparities in aging PWH. Moreover, our analysis highlights the critical role of demographic and structural determinants—such as race, age, and employment status—in shaping dietary behavior, emphasizing the need to embed nutrition-focused strategies into broader models of HIV care.

In our analysis, older age was independently associated with higher diet quality, as measured by the HEI-2015. This finding aligns with previous population-based studies suggesting that advanced age may be linked to increased adherence to dietary recommendations.^[34–36]^ For example, Dorrington et al.^[35]^ developed a Diet Quality Index specifically for older adults (DQI-65) and applied it to a sample of 871 UK adults aged 65 years and older. They reported that higher diet quality scores were associated with favorable health markers, including lower C-reactive protein levels and reduced BMI among older adults. Together these findings support the hypothesis that older individuals may adopt healthier dietary practices, possibly due to increased awareness of chronic disease management, regular clinical follow-up, or having more time and resources to dedicate to healthcare.

We also observed that Black participants had significantly lower HEI-2015 scores compared to White participants, underscoring racial disparities in diet quality. Similar findings were reported by Abdulai et al.^[37]^ in a cross-sectional study of 440 PWH in Ghana. Using the Individual Dietary Diversity Score as a proxy for diet quality, they found that only 14% of participants had a “good diet,” while 31% exhibited poor dietary diversity. Age was inversely associated with diet quality (AOR = 0.966, 95% CI: 0.936–0.997; *p* = 0.031), highlighting sociodemographic vulnerabilities. Although race was not directly examined, the study emphasizes the role of structural inequities, including food access and affordability, in shaping dietary behavior. Moreover, we found that the absence of current pain was marginally associated with higher HEI-2015 scores. These findings are consistent with recent research^[16]^ demonstrating that adherence to a high-quality diet—rich in fruits, vegetables, and whole grains—was associated with lower levels of chronic bodily pain in a cohort of 654 adults aged approximately 50 years, independent of age and body mass index. These data collectively illustrate that diet quality in PWH is closely tied to broader social determinants of health, including race, and physical symptom burden, which must be addressed in future interventions.

PHW in the highest tertile of HEI-2015 scores had markedly greater intake of dietary fiber, protein, and several key micronutrients—including magnesium, potassium, and vitamins D, E, K, and C. These participants also exhibited more favorable energy distribution across macronutrients and higher scores across most HEI-2015 components. These findings are consistent with those of Kirkpatrick et al. (2021),^[38]^ who compared observed versus reported dietary intake data in two feeding studies conducted in the U.S. (*n*=81 and *n*=302, aged 20–70). Their analysis demonstrated that HEI-2015 scores based on 24-hour dietary recalls are valid and reasonably accurate, supporting the reliability of our own dietary assessments. In the context of HIV, Malindisa et al.^[39]^ examined dietary patterns and inflammation in a cross-sectional study of 574 Tanzanian adults with and without HIV. Their results showed that adherence to a vegetable-poor dietary pattern was associated with elevated levels of fecal myeloperoxidase, a marker of intestinal inflammation. These findings reinforce the potential anti- inflammatory benefits of nutrient-dense diets and the value of HEI-2015 in capturing these differences.

This study has some limitations. First, its cross-sectional design precludes causal inference regarding the directionality of associations. However, we attempted to address this limitation by adjusting for a wide range of confounders, including clinical, behavioral, and social factors. Second, dietary data were self-reported, which may introduce recall bias; nonetheless, we minimized this concern by employing repeated 24-hour recalls administered by trained dietitians using a validated system. Among the study’s strengths are: (1) the inclusion of a large, geographically and racially representative sample of middle-aged and older PWH; (2) the use of standardized HEI-2015 scoring to quantify diet quality; (3) the incorporation of robust patient-reported outcomes and clinical covariates; and (4) the application of validated dietary assessment tools. These strengths enhance the generalizability and validity of our findings.

Clinically, our results support the integration of dietary screening and counseling into routine HIV care, particularly for older and racially marginalized individuals. Scientifically, this work contributes to the emerging literature on aging with HIV and offers a framework for identifying modifiable targets to improve nutrition-related outcomes. Future research should employ longitudinal designs to investigate causality and explore mechanisms by which social and structural factors shape diet quality over time.

## Conclusion

This study characterized the dietary patterns of predominantly middle-aged and older PWH and identified demographic and contextual correlates of diet quality. Individuals with higher HEI-2015 scores exhibit significantly improved nutrient intake. Our findings demonstrate that better diet quality in PWH is associated with older age, White race, full-time employment, and absence of pain. These results emphasize the importance of integrating nutritional support into HIV care models and highlight critical disparities that must be addressed to improve health equity in this aging population.

## Data Availability

All data produced in the present study are available upon reasonable request to the authors.

## REFERENCES

1. Oyetunji IO, Duncan A, Booley S, Harbron J. Diet quality, food insecurity and risk of cardiovascular diseases among adults living with HIV/AIDS: a scoping review protocol. BMJ Open 2021; 11(10):e047314.

2. Willig A, Wright L, Galvin TA. Practice Paper of the Academy of Nutrition and Dietetics: Nutrition Intervention and Human Immunodeficiency Virus Infection. J Acad Nutr Diet 2018; 118(3):486–498.

3. Collins LF, Palella FJ, Jr., Mehta CC, Holloway J, Stosor V, Lake JE, et al. Aging-Related Comorbidity Burden Among Women and Men With or At-Risk for HIV in the US, 2008-2019. JAMA Netw Open 2023; 6(8):e2327584.

4. Obare LM, Temu T, Mallal SA, Wanjalla CN. Inflammation in HIV and Its Impact on Atherosclerotic Cardiovascular Disease. Circ Res 2024; 134(11):1515–1545.

5. Yeung SSY, Kwan M, Woo J. Healthy Diet for Healthy Aging. Nutrients 2021; 13(12).

6. High KP, Brennan-Ing M, Clifford DB, Cohen MH, Currier J, Deeks SG, et al. HIV and aging: state of knowledge and areas of critical need for research. A report to the NIH Office of AIDS Research by the HIV and Aging Working Group. J Acquir Immune Defic Syndr 2012; 60 Suppl 1(Suppl 1):S1–18.

7. Krebs-Smith SM, Pannucci TE, Subar AF, Kirkpatrick SI, Lerman JL, Tooze JA, et al. Update of the Healthy Eating Index: HEI-2015. J Acad Nutr Diet 2018; 118(9):1591–1602.

8. Webel AR, Schexnayder J, Cioe PA, Zuniga JA. A Review of Chronic Comorbidities in Adults Living With HIV: State of the Science. J Assoc Nurses AIDS Care 2021; 32(3):322–346.

9. Qin XM, Allan R, Park JY, Kim SH, Joo CH. Impact of exercise training and diet therapy on the physical fitness, quality of life, and immune response of people living with HIV/AIDS: a randomized controlled trial. BMC Public Health 2024; 24(1):730.

10. Millar SR, Navarro P, Harrington JM, Perry IJ, Phillips CM. Dietary Quality Determined by the Healthy Eating Index-2015 and Biomarkers of Chronic Low-Grade Inflammation: A Cross-Sectional Analysis in Middle-to-Older Aged Adults. Nutrients 2021; 13(1).

11. Gherasim A, Arhire LI, Nita O, Popa AD, Graur M, Mihalache L. The relationship between lifestyle components and dietary patterns. Proc Nutr Soc 2020; 79(3):311–323.

12. Wetherill MS, Caywood LT, Hartwell ML, Bakhsh C, Weiser SD. Defining what Matters: Use of Q Methodology to Identify Food Values among People Living with HIV Affected by Food Insecurity. J Health Care Poor Underserved 2024; 35(4S):166–185.

13. Muhammad JN, Fernandez JR, Clay OJ, Saag MS, Overton ET, Willig AL. Associations of food insecurity and psychosocial measures with diet quality in adults aging with HIV. AIDS Care 2019; 31(5):554–562.

14. Odoms-Young A, Brown AGM, Agurs-Collins T, Glanz K. Food Insecurity, Neighborhood Food Environment, and Health Disparities: State of the Science, Research Gaps and Opportunities. Am J Clin Nutr 2024; 119(3):850–861.

15. Anema A, Vogenthaler N, Frongillo EA, Kadiyala S, Weiser SD. Food insecurity and HIV/AIDS: current knowledge, gaps, and research priorities. Curr HIV/AIDS Rep 2009; 6(4):224–231.

16. Ward SJ, Coates AM, Baldock KL, Stanford TE, Hill AM. Better diet quality is associated with reduced body pain in adults regardless of adiposity: Findings from the Whyalla Intergenerational Study of Health. Nutr Res 2024; 130:22–33.

17. Khan A, Boomer KB, Chiu Y-CJ, Bekele T, Murzin K, Kroch A, et al. Advancing our understanding of employment as a social determinant of health among people with HIV in Ontario, Canada. Journal of Public Health 2025.

18. Tan ECK, Eshetie TC, Gray SL, Marcum ZA. Dietary Supplement Use in Middle-aged and Older Adults. J Nutr Health Aging 2022; 26(2):133–138.

19. Kassis A, Fichot MC, Horcajada MN, Horstman AMH, Duncan P, Bergonzelli G, et al. Nutritional and lifestyle management of the aging journey: A narrative review. Front Nutr 2022; 9:1087505.

20. Webel AR, Long D, Rodriguez B, Davey CH, Buford TW, Crane HM, et al. The PROSPER-HIV Study: A Research Protocol to Examine Relationships Among Physical Activity, Diet Intake, and Symptoms in Adults Living With HIV. J Assoc Nurses AIDS Care 2020; 31(3):346–352.

21. von Elm E, Altman DG, Egger M, Pocock SJ, Gotzsche PC, Vandenbroucke JP, et al. The Strengthening the Reporting of Observational Studies in Epidemiology (STROBE) statement: guidelines for reporting observational studies. J Clin Epidemiol 2008; 61(4):344–349.

22. Kitahata MM, Rodriguez B, Haubrich R, Boswell S, Mathews WC, Lederman MM, et al. Cohort profile: the Centers for AIDS Research Network of Integrated Clinical Systems. Int J Epidemiol 2008; 37(5):948–955.

23. Moshfegh AJ, Rhodes DG, Baer DJ, Murayi T, Clemens JC, Rumpler WV, et al. The US Department of Agriculture Automated Multiple-Pass Method reduces bias in the collection of energy intakes. Am J Clin Nutr 2008; 88(2):324–332.

24. Basiotis P, Carlson A, Gerrior S, Juan W, Lino M. The Healthy Eating Index: 1999-2000. USDA, Center for Nutrition Policy and Promotion, CNPP-12. In. Washington, DC: USDA, Center for Nutrition Policy and Promotion, CNPP-12; 2002.

25. Justice AC, Holmes W, Gifford AL, Rabeneck L, Zackin R, Sinclair G, et al. Development and validation of a self-completed HIV symptom index. J Clin Epidemiol 2001; 54 Suppl 1:S77–90.

26. Herdman M, Gudex C, Lloyd A, Janssen M, Kind P, Parkin D, et al. Development and preliminary testing of the new five-level version of EQ-5D (EQ-5D-5L). Qual Life Res 2011; 20(10):1727–1736.

27. Kroenke K, Spitzer RL, Williams JB. The PHQ-9: validity of a brief depression severity measure. J Gen Intern Med 2001; 16(9):606–613.

28. Kroenke K, Wu J, Yu Z, Bair MJ, Kean J, Stump T, et al. Patient Health Questionnaire Anxiety and Depression Scale: Initial Validation in Three Clinical Trials. Psychosom Med 2016; 78(6):716–727.

29. Babor TF, Higgins-Biddle J, Saunders JB, Monteiro MG. AUDIT-The Alcohol Use Disorders Identification Test: Guidelines for Use in Primary Heath Care. Second Edition. Geneva: Geneva: World Health Organization; 2001.

30. Group WAW. The Alcohol, Smoking and Substance Involvement Screening Test (ASSIST): development, reliability and feasibility. Addiction 2002; 97(9):1183–1194.

31. Frampton CL, Hughes-Webb P. The Measurement of Pain. In: *Clinical Oncology*; 2011. pp. 381–386.

32. Young J, Jeganathan S, Houtzager L, Di Guilmi A, Purnomo J. A valid two-item food security questionnaire for screening HIV-1 infected patients in a clinical setting. Public Health Nutr 2009; 12(11):2129–2132.

33. U.S. Department of Agriculture, Food and Nutrition Service, Center for Nutrition Policy and Promotion. Average Healthy Eating Index-2020 Scores for the U.S. Population - Total Ages 2 and Older and by Age Groups, WWEIA, NHANES 2017-2018. In; 2023.

34. Drewnowski A, Henderson SA, Driscoll A, Rolls BJ. The Dietary Variety Score: assessing diet quality in healthy young and older adults. J Am Diet Assoc 1997; 97(3):266–271.

35. Dorrington N, Fallaize R, Hobbs D, Weech M, Lovegrove JA. Diet Quality Index for older adults (DQI-65): development and use in predicting adherence to dietary recommendations and health markers in the UK National Diet and Nutrition Survey. Br J Nutr 2022; 128(11):2193–2207.

36. Robinson SM, Westbury LD, Cooper R, Kuh D, Ward K, Syddall HE, et al. Adult Lifetime Diet Quality and Physical Performance in Older Age: Findings From a British Birth Cohort. *The journals of gerontology Series A*, Biological sciences and medical sciences 2017; 73(11):1532–1537.

37. Abdulai K, Torpey K, Kotoh AM, Laar A. Associated factors of diet quality among people living with HIV/AIDS in Ghana. BMC Nutr 2024; 10(1):90.

38. Kirkpatrick SI, Dodd KW, Potischman N, Zimmerman TP, Douglass D, Guenther PM, et al. Healthy Eating Index-2015 Scores Among Adults Based on Observed vs Recalled Dietary Intake. J Acad Nutr Diet 2021; 121(11):2233–2241 e2231.

39. Malindisa EK, Dika H, Rehman AM, Kweka B, Todd J, Olsen MF, et al. Associations between dietary patterns and intestinal inflammation among HIV-infected and uninfected adults: A cross-sectional study in Tanzania. PLoS One 2024; 19(12):e0311693.

